# Clinical Decision Support with a Comprehensive in-EHR Patient Tracking System Improves Genetic Testing Follow Up

**DOI:** 10.1101/2023.01.24.23284923

**Authors:** Ian M. Campbell, Dean J. Karavite, Morgan L. McManus, Fred C. Cusick, David C. Junod, Sarah E. Sheppard, Eli M. Lourie, Eric D. Shelov, Hakon Hakonarson, Anthony A. Luberti, Naveen Muthu, Robert W. Grundmeier

**Author notes:** **Address correspondence to:** Ian Campbell, MD PhD, Department of Biomedical and Health Informatics, Roberts Ctr Pediatric Research, 2716 South Street, Philadelphia, PA 19146. Unit on Vascular Malformations, Eunice Kennedy Shriver National Institute of Child Health and Human Development, Bethesda, MD, USA.

## Abstract

**Objective:** We sought to develop and evaluate an electronic health record (EHR) genetic testing tracking system to address the barriers and limitations of existing spreadsheet-based workarounds.

**Materials and Methods:** We evaluated the spreadsheet-based system using mixed effects logistic regression to identify factors associated with delayed follow up. These factors informed the design of an EHR-integrated genetic testing tracking system. After deployment we assessed the system in two ways. We analyzed EHR access logs and note data to assess patient outcomes and performed semi-structured interviews with users to identify impact of the system on work.

**Results:** We found that patient-reported race was a significant predictor of documented genetic testing follow up, indicating a possible inequity in care. We implemented a CDS system including a patient data capture form and management dashboard to facilitate important care tasks. The system significantly speeded review of results and significantly increased documentation of follow-up recommendations. Interviews with system users identified key team members ensuring success and revealed that the system addresses a number of sociotechnical factors that collectively result in safer and more efficient care.

**Discussion:** Our new tracking system ended decades of workarounds for identifying and communicating test results and improved clinical workflows. Interview participants related that the system decreased cognitive and time burden which allowed them to focus on direct patient interaction.

**Conclusion:** By assembling a multidisciplinary team, we designed a novel patient tracking system that improves genetic testing follow up. Similar approaches may be effective in other clinical settings.

## Introduction

Genetic testing can unlock the promise of precision medicine, potentially informing prognosis, management, surveillance, and recurrence risk [1]. However, to be useful, the testing must be reviewed and interpreted by professionals who can implement care based on the knowledge gained and inform the patient or their family members of its meaning. To facilitate such care, researchers have identified a range of considerations for incorporating genetics into the electronic health record (EHR) including data standards, data sharing, and the return or results process [2–4]. While management of genetic testing presents many unique challenges, such as insurance prior authorization, multiple specialized reference labs and long turn-around times, parallels can also be made to more general test result workflows. Various studies have demonstrated clinician dissatisfaction, inefficiencies, and safety problems in the EHR and in managing a broad range of clinical tests in internal medicine and primary care [5,6].

For example, one study found that three fourths of physicians did not routinely notify patients of normal test results and that up to one third of physicians did not always notify patients about abnormal test results [7]. The same study found that fewer than one fourth of physicians had a reliable method for identifying patients overdue for follow up of abnormal test results. A recent analysis of malpractice cases by a large insurer showed that about one quarter of diagnosis-related malpractice cases can be attributed to failures in the follow up system [5]. Taken together, these studies suggest that current test result follow up systems do not meet the needs of patients and physicians. When clinicians are not provided tools to support their work, they create their own tools and processes referred to as workarounds. These workarounds can be either paper-based or electronic (for example spreadsheets or external databases).

While studies do exist on the productivity of genetic counselors [8–11], there appears to be a gap in the literature regarding the specific task of results tracking. From our personal observations and comments of other genetics professionals, the most widely used EHRs lack comprehensive functionality to track genetic test results. Some genetic counselors in our study report that they were instructed in graduate school to use spreadsheets to manage result tracking. These spreadsheets qualify as an electronic workaround, but they have many limitations including being separate from the patient record; being susceptible to security issues, loss or being out of date; and frequently preventing multiple simultaneous users. There have been attempts to design more effective test result systems [12], including systems specifically addressing genetic testing [2]. These systems suffer from many of the same issues listed above for spreadsheet-based workarounds, and often require workflows outside the EHR. In a recent study, Scott and Martin implemented a genetic testing data collection system within a popular commercial EHR [2]. This facilitated centralized reporting in their practice, but the authors did not describe a comprehensive system for management of patients over time.

As an initial area to improve care, we chose to focus on inpatient genetic testing. At the Children’s Hospital of Philadelphia, like many academic pediatric hospitals, the inpatient clinical genetics evaluation workflow involves the primary care team’s attending physician requesting consultation of a geneticist (Figure 1). The geneticist evaluates the patient and makes recommendations for genetic testing. It is the responsibility of the primary care team to determine if the recommendations fit well with the overall treatment plan and then send samples to the hospital or outside reference laboratory for analysis. The laboratory issues a test report which is in turn entered into the patient’s EHR record. Because the genetic testing takes weeks or even months to complete and often includes more than one laboratory reporting results at different times, the patient may be discharged before any results become available.

**Figure 1.**
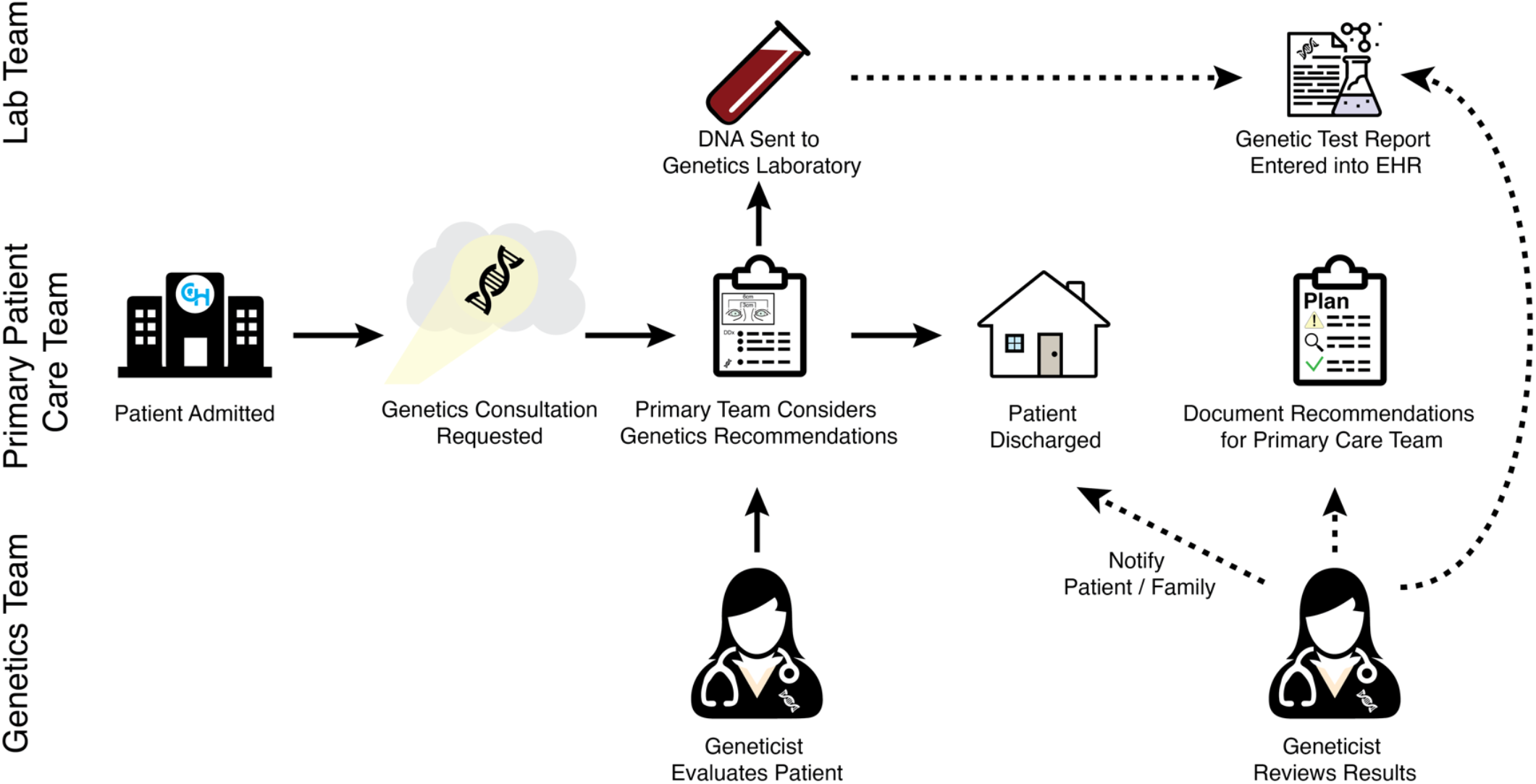
Swimlane diagram of the clinical genetics inpatient consultation workflow. Patients admitted to the hospital may be suspected by their primary inpatient care team of having an underlying genetic etiology for their medical problems. A consultation may be requested. The genetics team (including an attending physician) evaluates the patients and leaves recommendations for the primary care team. These recommendations often include genetic testing, which is then sent to the hospital genomics lab or an outside reference lab. Because genetic testing may take weeks or months, the patient may be discharged prior to testing resulting. Once testing is complete, the test report is entered into the patient’s EHR record. Ideally, the medical geneticist becomes aware of the result and interprets them in the context of the patient. The geneticist informs the patient or their family members and documents their recommendations in the EHR for other care team members.

The clinical genetics team should become aware of the results and interpret them based on patient’s presentation and family history and document their recommendations in the EHR. Additional challenges may hamper this workflow, including high demand for limited clinical resources, turnover of care teams, mismanaged paper documents, and manual result entry into the EHR. Historically, our clinical genetics team developed paper records or Microsoft Office^®^ documents as workarounds. The development of such artifacts as “workarounds” to the EHR is a practice not uncommon in other medical contexts [13–15]. Despite implementing these types of workarounds in our own health system, we found anecdotally that results sometimes go unnoticed (Figure 2).

**Figure 2.**
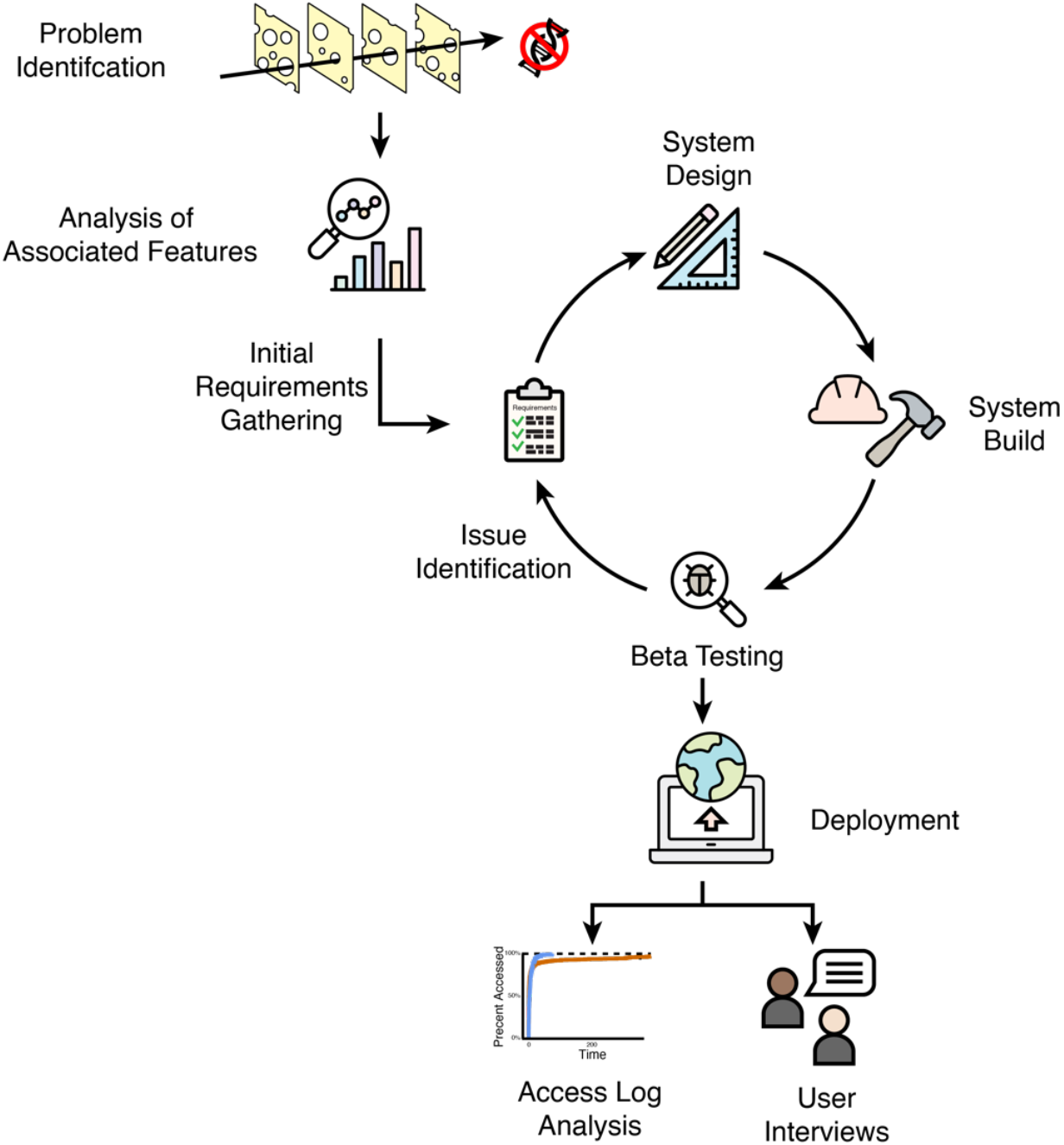
Project overview. We became aware of anecdotal reports of results going unnoticed. To better understand the problem, we performed an analysis of patient characteristics that were associated with the presence or absence of documented follow up recommendations. Guided by this analysis, we gathered initial requirements for a successful system to address these issues. We adopted an agile project development lifecycle and designed the system, built and deployed the system, and initiated a limited clinical beta test. Based on our testing, we identified issues and iterated on the system. We then provided user education and deployed the system to the entire inpatient service. Following implementation, we analyzed patient outcomes and conducted semi-structured interviews of users.

Researchers in other health systems have reported attempts at improving these workflows by implementing data collection directly in the EHR, but ultimately users reportedly export the data outside the EHR into spreadsheet programs for further management [2]. Our objective was to improve genetic testing result interpretation and documentation of follow up recommendations. Specifically, we (1) sought to understand the factors potentially associated with follow up, (2) developed a clinical decision support (CDS) mechanism using a comprehensive in-EHR tracking system. (3) assessed the effectiveness of our intervention using access logs and EHR documentation, and (4) interviewed end users of the system to understand factors key to successful implementation.

## Materials and Methods

The Institutional Review Board at Children’s Hospital of Philadelphia determined that this study met exemption criteria under 45 CFR 46.104(d)4(iii) as protocol 20-018111. A waiver of HIPAA authorization under 45 CFR 164.512(i)(2)(ii) was granted to access identifiable information from the medical records. Statistical analyses and data visualization were performed with the R statistical programming language. Multivariate mixed-effects logistic regression analyses were performed with the GLMMadaptive package to facilitate efficient calculation of marginal effects conditional on the average patient. Survival analysis was performed using the survminer package.

### Setting and Participants

This study was performed at the Children’s Hospital of Philadelphia, a pediatric quaternary referral hospital. The Section of Clinical Genetics cares for approximately 5,500 children per year with suspected or confirmed genetic disease in the ambulatory setting. Additionally, the section maintains an inpatient consultation service which provides evaluation and management recommendations for approximately 800 children per year. This manuscript focuses on the inpatient consultation workflow. At the time of the study, the section included 10 attending physicians, 11 residents or fellows, and 3 genetic counselors. Clinician participants for system development and user interviews were recruited from the section. Trainees from other divisions such as Child Neurology and Developmental Pediatrics were involved in a small minority of consultations.

### Analysis of Factors Associated with Results Follow Up

To understand factors associated with results follow up, we accessed EHR data for children undergoing inpatient genetic consultation during the 1 year prior to implementation of the tracking system. We collected demographic information about the participants including sex, age at time of consultation, gestational age, self-reported race and ethnicity, and home address census tract. From the hospital admission we collected information about the care team, genetic testing performed, and documentation of follow up recommendations.

### System Development and Deployment

We examined the various systems employed by our division for patient tracking over the previous 10-year period. We undertook informal interviews of users of these systems both over time and fulfilling various roles. From these interviews, we identified strengths and limitations of these systems to inform the design requirements of a user-centered EHR-based system. We adopted an Agile project development approach (Figure 2). Early designs were reviewed with primary users of the system and modified based on their feedback. During the design process, we identified a number of critical design requirements that could not be addressed by base EHR functionalities. To deal with these, we co-opted functionalities originally developed by our EHR vendor for disparate tasks and combined them in novel ways to create functionality not provided by the standard EHR. To overcome other obstacles, we were forced to program custom low-level functions to enable key functionality.

The initial system was beta tested in a limited clinical environment before deployment to the entire inpatient service. During this process, we identified and addressed issues which included technical, workflow and usability problems. We next developed and delivered training to end users. After deployment, based on feedback from users, we made rapid iterative changes to the system.

### System Outcomes

To assess performance before and after implementation of our tracking system, we identified two outcome measures: (1) time from result to access by genetic team member, and (2) time to follow up recommendation documented in a clinical note. We collected information about clinical encounters, note documentation, and genetic testing from the EHR. Because the time of result review is not consistently recorded by the EHR, we used detailed access logs—the record of all user actions throughout the system. We first created a list of all members of the genetics team (attending, resident, fellow and genetic counselors). We next identified log entry types as a proxy which could reasonably indicate that the user reviewed genetic testing results (for example “results review accessed” or “results table viewed”). Although we cannot be certain that the user actually viewed the genetic test result, we believe this is a reasonable proxy as the genetics team would have little reason to be accessing the patient’s results other than to view the genetic test. Likewise, we would expect that errors should occur in both the control and intervention groups equally.

### Post Deployment User Interviews

We developed a semi-structured interview guide to elicit open-ended responses from users of the tracking tool. To capture information on the broader work domain, interview questions were guided by the sociotechnical model, Safety Engineering in Patient Safety (SEIPS) 2.0 [16]. We recruited a sample of genetic counselors, residents, fellows, and attending physicians in the inpatient genetics section. Participants provided informed consent and were then interviewed individually using web conferencing software where the session was audio recorded and then machine transcribed. Two study team members deductively analyzed the transcripts using themes based on components of SEIPS 2.0. An initial review of transcripts was performed where results were compared, and consensus reached on thematic analysis.

## Results

### Factors Associated with Results Follow up

To inform our development of a more successful system, we analyzed the outcomes of 768 inpatient genetics consultations during the 1-year period prior to implementation of the tracking system. 58% of patients had no subsequent documentation from a provider or staff member that included clinical genetics. Of those patients, 78% (or 45% of the total), had genetic testing performed after their consultation but still had no follow up documented by the genetics team. It is possible that some of these patients had results reviewed and documented by their primary team. We worried that limitations in our follow up infrastructure perpetuated inequities in the delivery of care to patients with historically marginalized backgrounds. To this end, we performed an analysis of factors associated with documentation of follow up recommendations using a mixed-effects logistic regression (Table 1). Unsurprisingly, we found that genetic testing was strongly significantly associated with documentation of follow up recommendations. We also found that patient reported race was significantly associated with follow up documentation (p = 0.011, Wald test), with patients reporting non-Hispanic Black background less likely to have recommendations documented (raw 40.4% versus 42.1% for all other groups, adjusted marginal odds ratio 0.82). These analyses validated our concerns about inequities.

**Table 1:** Mixed Effects Logistic Regression Predictors of Follow Up Documentation

| Predictor | Category | Odds Ratio | p-value |
| --- | --- | --- | --- |
| Any Genetic Testing | No | *Reference* | 0.0008 |
|  | Yes | 2.01 |  |
| Sex | Female | *Reference* | 0.1335 |
|  | Male | 0.81 |  |
| Hospital Length of Stay (Weeks) |  | 1.03 | < 0.0001 |
| Admission Type | Born in Hospital | *Reference* | 0.0221 |
|  | Other | 1.43 |  |
| Preferred Language | English | *Reference* | 0.2736 |
|  | Other | 0.73 |  |
| Race | Non-Hispanic White | *Reference* | 0.0114 |
|  | Hispanic or Latino | 1.31 |  |
|  | Non-Hispanic Black | 0.82 |  |
|  | Other | 1.00 |  |
| Payor | Commercial | *Reference* | 0.2154 |
|  | Government | 0.78 |  |
|  | Charity | 0.32 |  |
| Observations: 768 |  |  |  |

### System Design

A review of our current workflow revealed that, for over a decade, our division relied on tables contained within Microsoft Word® documents housed on network share drives to keep track of inpatient consults. To review the patient test results, the system involved manually copying information out of the EHR. This was a process that was extremely time consuming, caused conflicts with multiple users attempting to access the document simultaneously, and was not ideal from a security perspective. We anecdotally heard patient stories of results going unnoticed and not being returned to families, that were only discovered when the family or other care provider followed up, often months later. As an initial attempt at addressing these issues, we created a database in REDCap to track patients we had previously evaluated [17]. However, we found that it was not consistently used by providers and suffered many of the same inefficiencies of the word processing documents (particularly the need to duplicate information).

Building upon our previous attempts and statistical analysis, we developed a tracking system with two main components. Part 1 is a data capture instrument which is available to clinicians in note templates and from patient lists (Figure 3). The capture form is optimized to reduce double documentation and provides customized suggestions but gives the clinician an opportunity to alter the parameters based on clinical judgement. Part 2 is a management dashboard which gives clinicians an overview of patients with outstanding tasks, provides easy access to information required to make decisions, and facilitates communication with the primary team and the patient’s family (Figure 4). One key clinical decision support tool is the inclusion of icons to clearly delineate the patient’s status in the diagnostic process. These include “testing recommended but not yet in process,” “testing in process” and “results to return.” The system automatically displays “overdue” icons based on standard estimates of test turnaround times. The user can easily sort or filter the list to view only their patients or those with overdue tasks to follow up.

**Figure 3.**
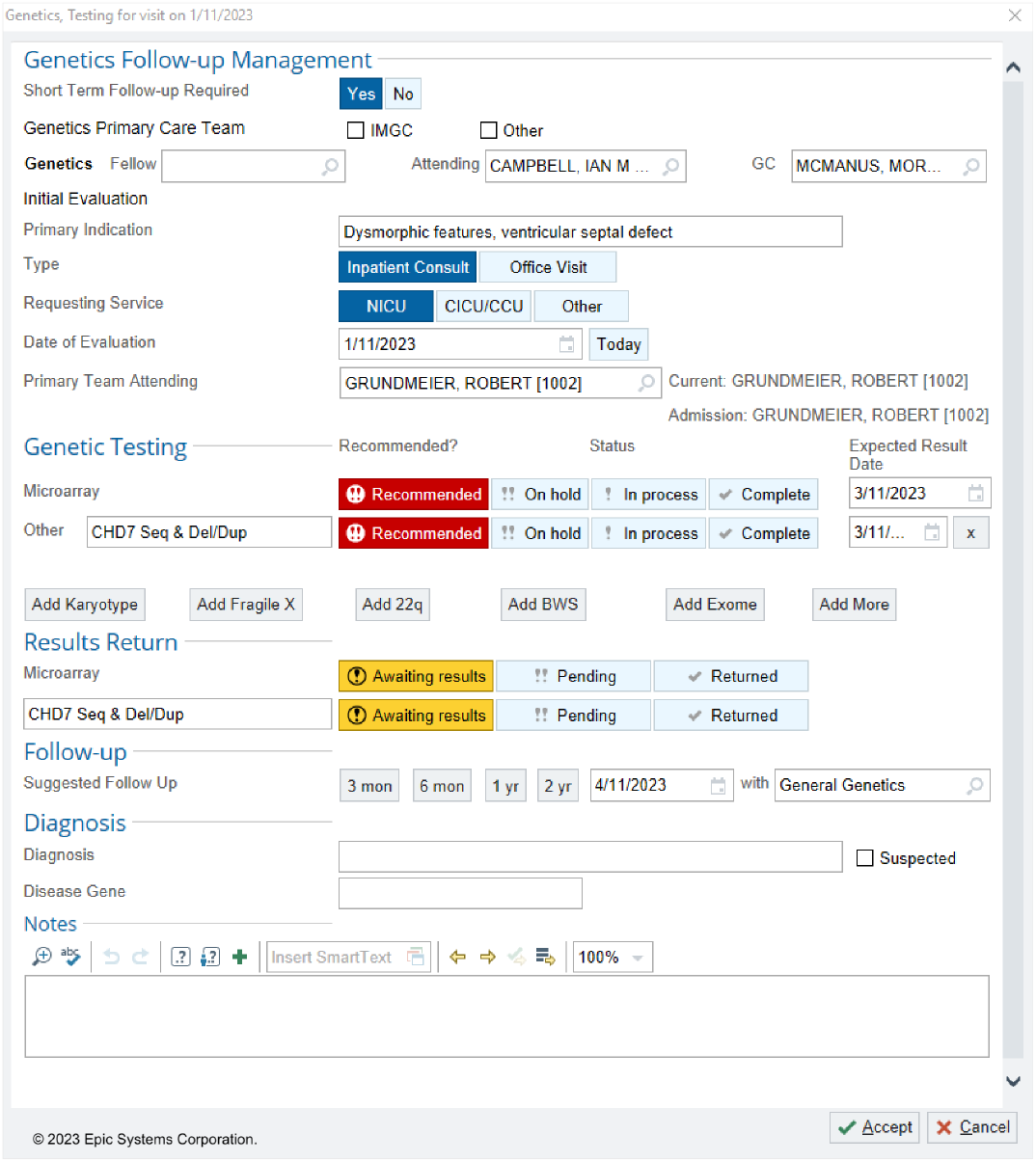
Data capture instrument for the in-EHR tracking system. We created a data capture form based on our EHR vendor’s standard functionality (a SmartForm™). The system automatically enters data by pulling it from elsewhere in the system. For example, “Inpatient Consult” is automatically selected if the patient is currently admitted. However, clinicians may override results. Clinicians are then asked to fill in information which cannot be readily ascertained from other sources. The data is stored in discrete fields for later use in reporting and documentation. All patient data presented in this figure is synthetic.

**Figure 4.**
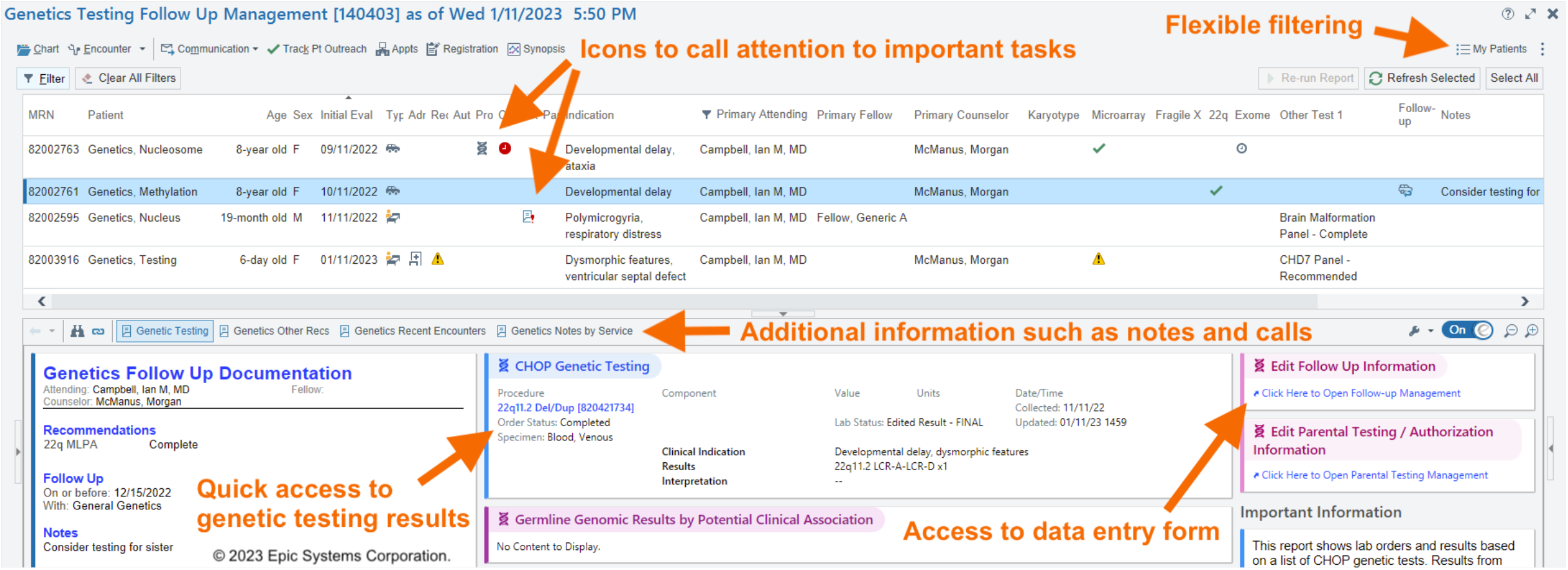
Management dashboard for the in-EHR tracking system. The dashboard is implemented using the vendor’s standard functionality (a Reporting Workbench™ Report). The top half of the management dashboard is a patient list which provides a high-level overview of the patient’s status within their genetic diagnostic odyssey. The standard functionality provides powerful filtering options to allow the clinician to, for example, only view their own patients by default. Icons provide key information about patient status. The bottom half of the dashboard provides additional details about the selected patient, including their genetic test results and recent communications with the patient’s family. There is also access to the data entry form (Figure 3) to document changes in patient status. All patient data presented in this figure is synthetic.

As part of deployment of the system, we delivered user education through a virtual meeting demonstrating the expected workflow and allowed users to ask questions. The development team also met individually with the users to answer questions and to receive feedback on barriers to work and system deficiencies. We created a “Job Aid” document which details use of the system as well as provides technical details about how the system actually functions.

### System Outcomes

Uptake of the tracking system by the user base was swift. In the first 6 months, 333 of 373 potentially eligible inpatient consults (89.3%) were entered into the system. The system facilitated multiple diagnoses that would have otherwise been missed, including a case of 22q11.2 deletion syndrome because genetic testing was inadvertently not sent by the primary management team. Considering patients evaluated in the first 6 months of deployment of the system, 171 of 275 (62.2%) inpatient consults who had genetic testing and were tracked by the system had follow up documentation within 60 days (Figure 5). This is a significantly greater proportion than those not tracked by the system over the previous year, 276 of 530 (52.1%, p < 0.005, Two-sided Fisher’s Exact Test). Although documentation rates differed among individual attending physicians, use of the tracking system was associated with increased follow up documentation by all but one provider. However, due to the small number of consults seen by the single provider with decreased rates (n = 8), the decrease is potentially due to randomness.

**Figure 5.**
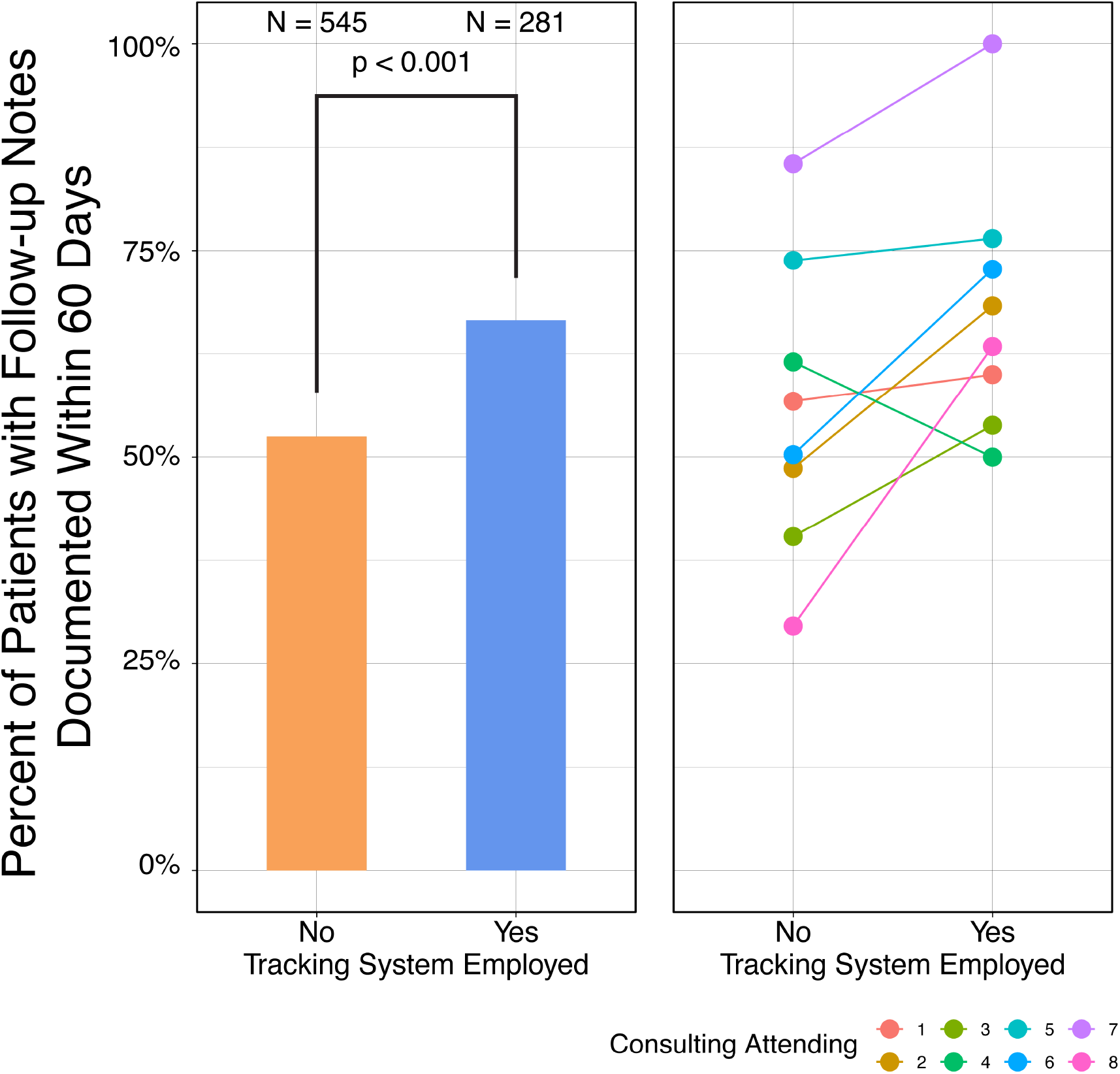
Changes in documentation of follow-up recommendations following system implementation. The left panel shows the percentage of patients who underwent genetic testing who had subsequent documentation of recommendations by genetics in the 60 days immediately following the results becoming available. The bar graph compare patients where the tracking system was not used (orange) to those where it was used (blue) in the 12 months preceding and 6 months following the implementation of the CDS tracking system. The numbers of patients were compared with a Fisher’s exact test. The left panel shows the same data but for each of the 8 consulting attending physicians who evaluated patients during both periods. Consulting attending number 4 (green dots and line) who decreased documented follow up percentage consulted on only 8 patients total, indicating possible stochasticity rather than an effect of the system itself.

Using EHR access log data, we found that review of genetic results was completed by a clinical genetics provider within one month for a great majority of patients regardless of tracking system utilization (Figure 6). However, the system helped facilitate review of small subset of patients that may otherwise have gone unnoticed (p < 0.001, Kolmogorov-Smirnov test). Indeed, all results were reviewed within 3 months (max 73 days) with the tracking system, whereas 8.2% of results remain un-accessed by that time without the system.

**Figure 6.**
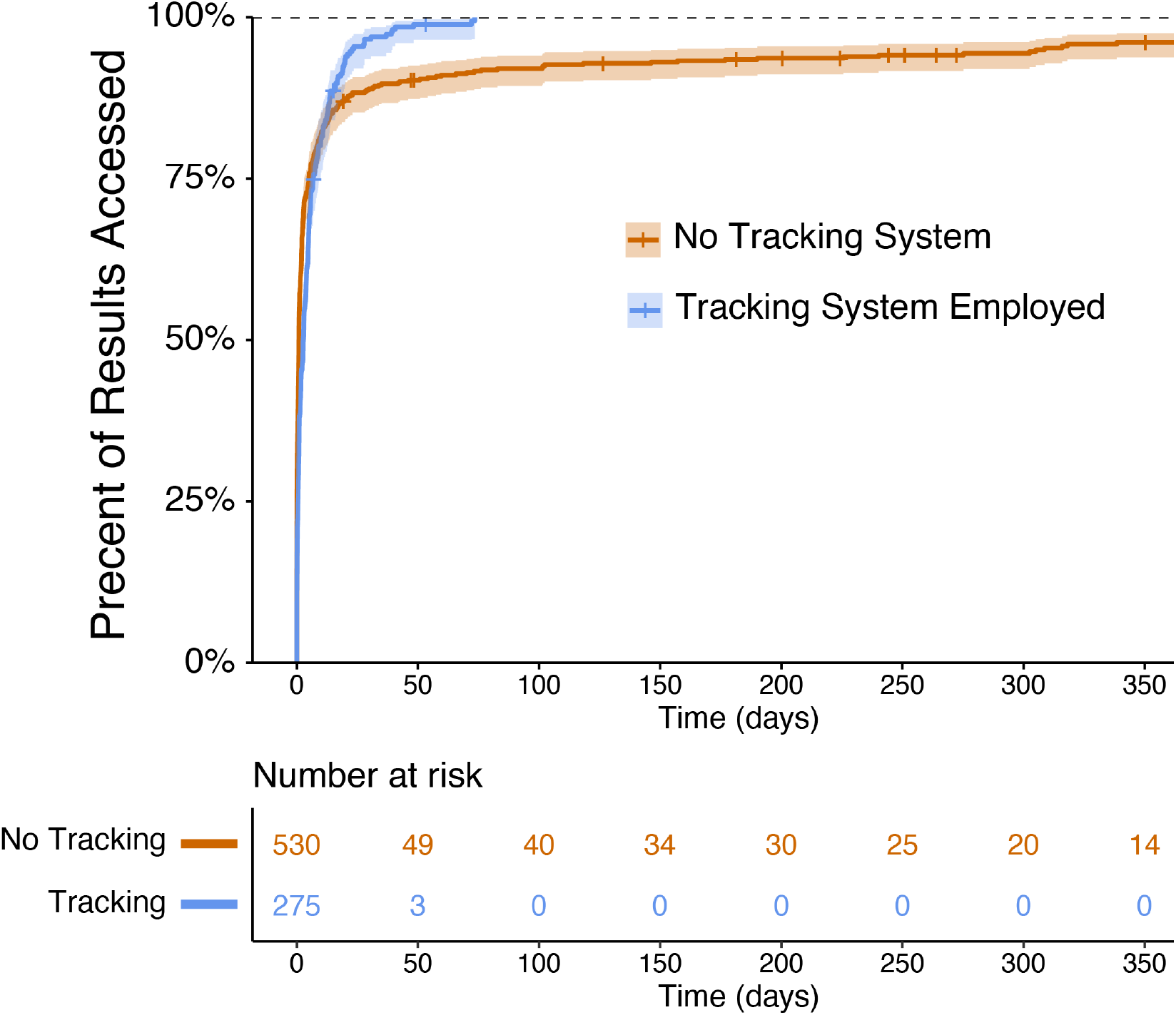
Survival analysis of time until genetic testing result review. The top panel shows the percentage of patients whose EHR lab results sections were reviewed by a genetics team member (genetic counselor, genetics resident, or genetics attending physician) over time for patients who were not tracked by the system (orange) compared to those who were tracked by the system (blue). Crosses indicate patients that were censored because their genetic testing had only been resulted for that long at the time of analysis. The bottom panel indicates the number of patients at risk.

### Post Deployment User Interviews

We interviewed a sample of 6 clinicians from the genetics department: (3) genetic counselors, (1) genetics resident physician, and (2) attending physicians. Interview transcripts were deductively analyzed using components of the SEIPS model (Table 2) [16]. Genetic counselors reported being the primary users of the system, managing the results tracking of all patients and responding to any changes, issues, or results with individual patients and coordinating a response with members of the clinical team. Physicians reported being secondary users whose work is typically impacted indirectly by the system, though they also interact with the system directly to review their patients or assess practice patterns.

**Table 2.** SEIPS 2.0 components, themes identified in our interviews and illustrative quotes.

| SEIPS Component | Concept | Example User Quote |
| --- | --- | --- |
| Persons | Responsible primary user<br>Technically inclined champion | “I think the system that we've put together has been effective partially because there is a genetic counselor who provides continuity.”<br>“I think somebody trained in workflow analysis and then with a deep technical knowledge of the vendor’s system would be required to replicate the tracking system.” |
| Tools & Technology | Collaborative single source<br>Access to information, documentation and communication in one place | “[We] had this Excel sheet that only one person could edit at a time and no one knew why there were 18 colors.”<br>“Right now it's like running on duct tape Excel sheets that cannot be sustained if testing increases by even double, right?”<br>“One of the big problems is that the things you need to do are spread out all over the EHR, like you need to look at the lab results and you need to write a note and you need to communicate with the team. Those things were all spread out in different places.” |
| Tasks | Improves task time<br>Service needs to be aware of unimplemented recommendations<br>Users have situational awareness of result status | “I genuinely don't know how we kept track of patients before this system.”<br>“Because [the system allows us to check] the admitted patients daily, we were able to see that the recommended testing was actually cancelled and was never collected. So we called the primary team to put in the orders again. And then eventually it did come back positive, and we were able to make that diagnosis.” |
| Organization | Extremely large volume of patients evaluated in academic medical center | “One of the columns in [the previous workaround was] a little checkbox for every task that we recommended for the patient, and if we were to look at those, we'd see that like 50% at least of the things don't have an X through them because there was no bandwidth to go back and check literally hundreds of patients.” |
| External Environment | Insurance authorization delays testing | "I would say it takes 5 to 7 messages per patient to get an insurance authorization." |
| Adaptation | Unanticipated use as an educational tool<br>Users implement filters to facilitate a range of personalized tasks | "[I use it to] look back at what other people have done for similar problems ... if they had a similar presenting problem and I was kind of wavering on ... what to do, it can help guide management for current patients based on what previous attendings have done." |
| Professional Work | Communication between genetic counselors, geneticists, and referring providers<br>Plan and prioritize daily work<br>Enables a level of review not possible without the system | "I think there is sometimes miscommunication between our team and the primary team about genetic testing."<br>"You feel it in your heart when you come across results and you're paging through the EMR asking 'where is the telephone note [from where someone] told this patient about the results?' and you don't find it."<br>"This tracking system potentially has the opportunity to decrease burnout." |
| Collaborative Work | Plan family meetings to discuss recent results<br>Can help collaboration with families to return forms and samples | "I cannot tell you how many saliva kits are out there sitting on people's kitchen tables under a pile of mail that have never been returned." |
| Outcomes | No patient falls through the cracks<br>Clearly communicate our recommendations | "Since I started using [the system], I have felt a sense of safety ... and sleeping without waking up in the middle of the night thinking about little Johnny and if his test result is back." |

In discussing the previous system, participants had myriad comments on the barriers and limitations of spreadsheet-based workarounds. Genetic counselor participants described how their graduate school training (at different universities) included the use of spreadsheets to track results and how spreadsheet use was widespread in their clinical experience in other organizations. The spreadsheet-based systems were described as highly manual; prone to missing and incorrect data; difficult to share, version track and update; imposed cumbersome workloads including double documentation tasks, stress, and other inefficiencies; and perhaps most importantly created a single point of failure and patient safety hazard. Overall, genetic counselor participants described the previous system as a source of burnout and job dissatisfaction, taking time and attention away from important patient work. One participant noted, “I think it’s why genetic counselors are so frequently going to industry, because [in clinical practice there are many] patients that need things and that you’re worried they’re going to slip through the cracks because there is no way to track them.”

Genetic counselor participants described the new system as having a significant impact on their work by improving efficiency, confidence and trust in the information; communicating information and collaboration leading to distributed responsibility among the clinical team to improve safety; and improving job and professional satisfaction by freeing up time and effort for more meaningful and impactful patient work. Participants provided many examples of how the tracking tool facilitated communication and collaboration including between genetic counselors and across the clinical genetics team. Moreover, it also facilitated communication with the rest of the patient’s care team outside genetics, such as the inpatient treatment team or their primary care pediatrician.

Both genetic counselor and physician participants described several unanticipated positive adaptations of the tracking system. Genetic counselors described how the centralized sortable and filterable display of all patients with testing allowed them to go beyond managing results and to anticipate and plan work for the upcoming week. A resident physician participant described using the system as an informal teaching tool such that searching for patients with a similar diagnosis could inform possible evaluation courses for other patients. An attending physician participant described how they used the system to understand their own practice patterns.

## Discussion

Overall, we have implemented an in-EHR tracking system to serve as a CDS tool that facilitates genetic testing and follow up, improving both patient care and patient safety. The tool was quickly adopted by the clinical team. Our data suggests that the ability to easily see an overview of outstanding care tasks may expedite review of results and documentation of interpretation and recommendations. Interviews with users of the system also revealed some surprising results. First, we learned from multiple participants that the creation of workarounds external to the EHR such as Microsoft Excel® was recommended at genetic counseling training programs. Thus, it seems likely that such workarounds are in use across health systems, although additional study is needed. Second, participants realized the limitations of the previous workflow and how that affected their ability to provide proper patient care prior to the implementation of the system. The potential existence of patients with unreviewed genetic testing that might significantly influence their care was a major concern. Participants expressed appreciation that the system acts as a safety net to help prevent patients from slipping through the cracks. Some felt that this functionality of the system may help with burnout. This may become a more critical issue to address if genetic testing continues to become a larger component of healthcare.

While our analysis and interviews suggest that our tracking system can improve care, our approach required significant investment. First, implementation of the system required many hundreds of person-hours across a highly invested multidisciplinary team. We assembled key stakeholders from genetic counseling, clinical genetics, clinical informatics, and information services. Second, while most of the key features of our system were built using the vendor’s standard functionality, some of the time-saving features required highly complicated build or creation of custom code. This required inclusion of team members with substantial technical expertise and a robust system for documentation and testing. These technical dependencies may also complicate long-term maintenance of the tracking system as the vendor makes changes to the EHR and individual team members with technical knowledge leave the institution. Despite these issues which may hamper development in a smaller hospital setting, we are making this tool available to the broader community.

Moreover, our study was conducted in a single health system which may limit generalizability of results. Only a small number of genetic counselors and physicians were available for interview so their perspectives may not be reflective of individuals in these roles at other health systems. The improvement in follow up we observed in our pre-post evaluation could be susceptible to secular trends, but there were no other concurrent efforts to improve genetic test result follow up to suggest such trends may have affected our results.

Our initial review demonstrated health inequity. Specifically, patients reporting non-Hispanic black background were less likely to have recommendations documented in the EHR. Due to the small number of patients, we lacked power to determine if our system was able to address inequities in the delivery of genetic care that we discovered. This will be the focus of continued research.

Although we studied and addressed a specific clinical genetics and genetic testing workflow in one academic hospital, we suspect that the approach is highly relevant to other clinical settings. We have heard from clinicians at more than 10 other children’s hospitals that they also use Excel spreadsheets to track their patients (personal communication). We have also received feedback of workarounds in other pediatric specialties as well as in our previous study of workarounds used to facilitate palivizumab dosing in former premature infants [18]. While our tracking system is highly customized to particularities of the clinical genetic and genetic testing workflows, we suspect that a slightly modified approach could be used to track patients undergoing other evaluations requiring multiple tests to track often with longer follow up times, for example, neurology or rheumatology evaluation. Furthermore, our approach can be applied to multidisciplinary hospital teams, for example vascular anomaly programs, to facilitate the nurse coordinator workflow of tracking multiple tests among specialties.

In conclusion, we identified results review and follow up as a gap in genetics care that persisted despite extra-EHR workarounds including spreadsheets and custom databases. We assembled a multidisciplinary team and created a comprehensive in-EHR tracking system optimized to limit double documentation and to improve efficiency by providing rapid access to needed patient information. We found uptake of the system was swift and facilitated significantly improved review and return of patient results with the perception of marked improvement in patient care. Additional work will be required to ease implementation at other clinical sites, to apply this system to other programs, and to understand if this work can address health inequity.

## Data Availability

All data produced in the present study are available upon reasonable request to the authors

---

**Supplemental Figure 1.**
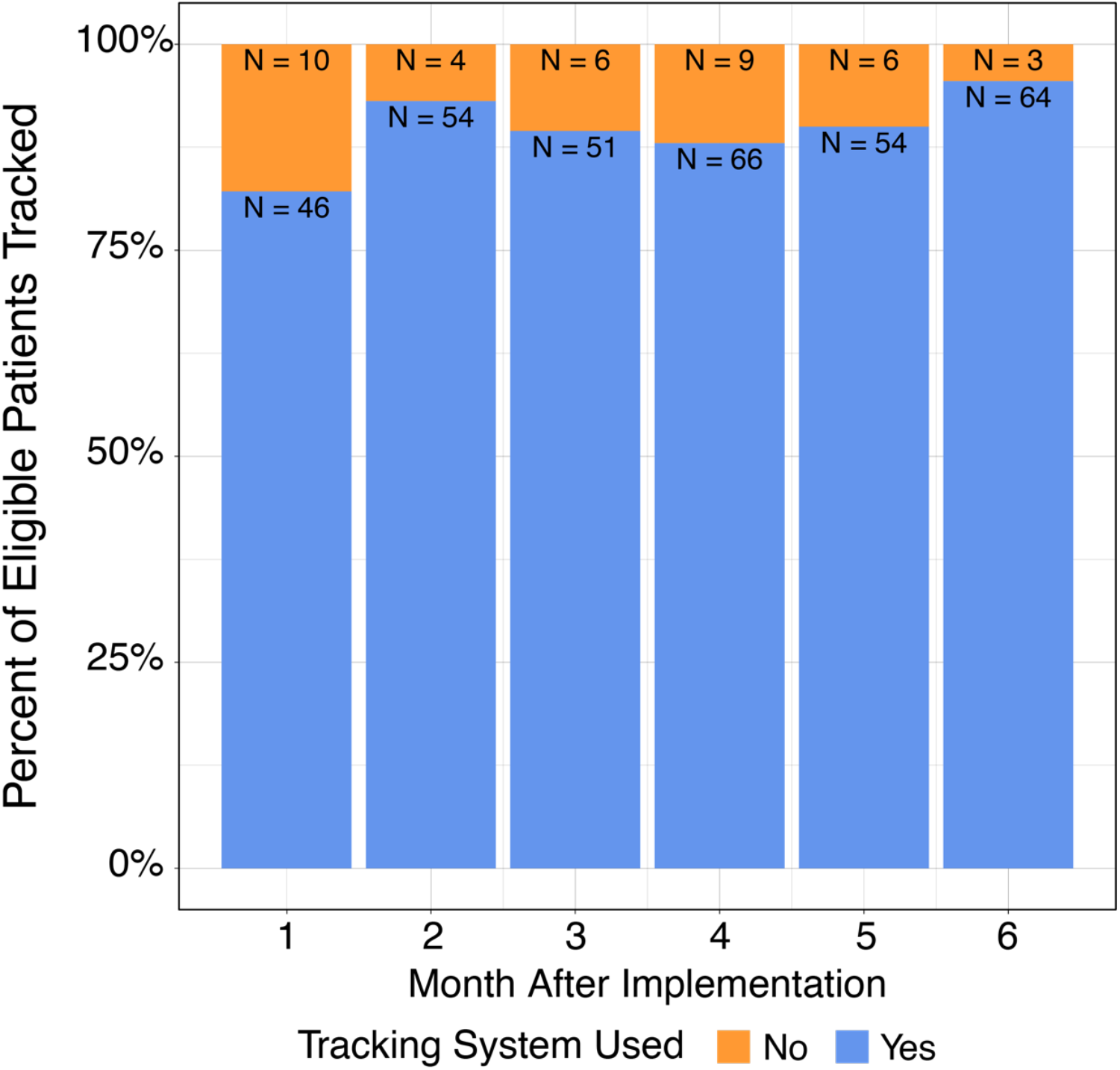
Uptake of the genetic tracking system after implementation. The bars indicate the percentage of inpatient consultations where the tracking system was employed by their care team. Blue indicates that the system was used and orange indicates that the system was not used.

